# Genome-wide Association Study of Pulmonary Function in Europeans and Africans from the UK Biobank Identifies Distinct Variants

**DOI:** 10.1101/2022.01.07.22268836

**Authors:** Musalula Sinkala, Samar S. M. Elsheikh, Mamana Mbiyavanga, Joshua Cullinan, Nicola J. Mulder

**Affiliations:** University of Cape Town, Faculty of Health Sciences, Institute of Infectious Disease and Molecular Medicine, Computational Biology Division, Anzio Rd, Observatory, 7925, Cape Town, South Africa; Pharmacogenetics Research Clinic, Campbell Family Mental Health Research Institute, Centre for Addiction and Mental Health, Toronto, ON, Canada

## Abstract

Pulmonary function is an indicator of well-being, and pulmonary pathologies are the third major cause of death worldwide. FEV1, FVC, and PEF are quantitively used to assess pulmonary function. We conducted a genome-wide association analysis of pulmonary function in 383,471 individuals of European and 5,978 African descent represented in the UK Biobank. Here, we report 817 variants in Europeans and 3 in Africans associated (p-values < 5 × 10^−8^) with three pulmonary function parameters; FEV1, FVC and PEF. In addition to 377 variants in Europeans previously reported to be associated with phenotypes related to pulmonary function, we identified 330 novel loci, including an *ISX* intergenic variant rs369476290 on chromosome 22 in Africans and a *KDM2A* intron variant rs12790261 on chromosome 11 in Europeans. Remarkably, we find no shared variants among Africans and Europeans. Enrichment analyses of variants separately for each ancestry background revealed significant enrichment for terms related to pulmonary phenotypes in Europeans but not Africans. Further analysis of studies of pulmonary phenotypes revealed individuals of European background are disproportionally overrepresented in datasets compared to Africans, with the gap widening over the past five years. Our findings offer a better understanding of the different variants that modify pulmonary function in Africans and Europeans, a significant finding for future GWAS studies and medicine.

## Introduction

Pulmonary function measures using the spirometer are indicators of respiratory health and predict morbidity and mortality^1,2^. These parameters, which include the force expiratory volume in 1-second (FEV1), forced vital capacity (FVC), and peak expiratory capacity (PEF), vary significantly among populations of different ancestry backgrounds^3^ and show strong evidence of genetic and environmental influences^2,4^.

During the last decade, large-scale genome-wide association studies (GWASs) have used various pulmonary parameters to evaluate the genomic loci associated with pulmonary function and related traits that have yielded hundreds of associated variants ^5–10^. These and other studies indicate that genomic loci associated with pulmonary function overlap with chronic obstructive pulmonary disease (COPD), asthma, pulmonary fibrosis, lung cancer, and other pulmonary phenotypes^1,8–10^. A recent GWAS based on the UK Biobank cohort (N = 50,008), including heavy smokers and never smokers, identified six loci associated with low FEV1^10^. Another study in individuals (N = 48,943) sampled from the extremes of pulmonary function distribution in UK Biobank identified 95 variants strongly associated with COPD susceptibility^8^. Importantly, these previous studies have applied the analyses on a selected population group of primarily European ancestry.

The UK Biobank cohort provides data on over 500,000 individuals that offers new prospects to identify variants associated with pulmonary function among Europeans and Africans using GWAS approaches by allowing for large-scale comparisons of lung function parameters^11^. Furthermore, by integrating the genetic association of FEV1, PEF, and FVC, a list of shared loci that collectively modify pulmonary function could be identified. We hypothesise that different genetic variants are associated with pulmonary function in Africans. Thus, their identification will provide additional information relevant to understanding pulmonary function in physiology and disease in district populations. However, to our knowledge, no GWAS study has been performed to compare the SNPs associated with the full range of FEV1, FVC and PEF parameters across the entire UK Biobank cohort and separately among Africans and Europeans.

Here, we compare variations in pulmonary function parameters among individuals of African and European ancestry represented in the UK biobank. First, we used the genome-wide associated summary statistics for three UK Biobank defined continuous pulmonary function parameters; FEV1, FVC, and PEF. Then, we conducted further analyses to identify genes, regions, and gene sets associated with each pulmonary phenotype. Furthermore, we evaluate the candidate phenotype loci in relation to published GWAS results. Overall, this approach allows us to report credible loci associated with pulmonary function among Africans and Europeans, which were enriched across many plausible genes and gene sets involved in pulmonary function or related phenotypes.

## Results

### UK Biobank pulmonary function demographics

There were 389,449 participants comprising of Europeans (N = 383,471) and Africans (N = 5,978). The mean age of participants at recruitment was 56.8 (standard deviation [Std] = 8.0) years and 51 (Std = 7.9) years for the Europeans and Africans, respectively.

### Lung function parameters vary between individuals of Europeans and African ancestry

We assessed the mean FVC, FEV1, and PEF between Europeans (N = 383,471) and Africans (N = 5,978) represented in the UK Biobank datasets. We found that the mean FVC was significantly higher in the Europeans (mean = 3.73 L) compared to the Africans (mean = 2.95 L), (Welch test: *t* = 48.35, p < 1 × 10^−320^; Figure 1a). Furthermore, we found that the FEV1 and the PEF were both significantly higher in Europeans (mean FEV1 = 2.82 L, mean PEF = 389.6 L/min) than those measured in the Africans (mean FEV1 = 2.28 L, mean PEF = 332.7 L/min), FEV1; *t* = 42.60, p = 1.0 × 10^−291^ (Figure 1b) and PEV; *t* = 24.06, p = 1.7 × 10^−107^; Figure 1c.

**Figure 1:**
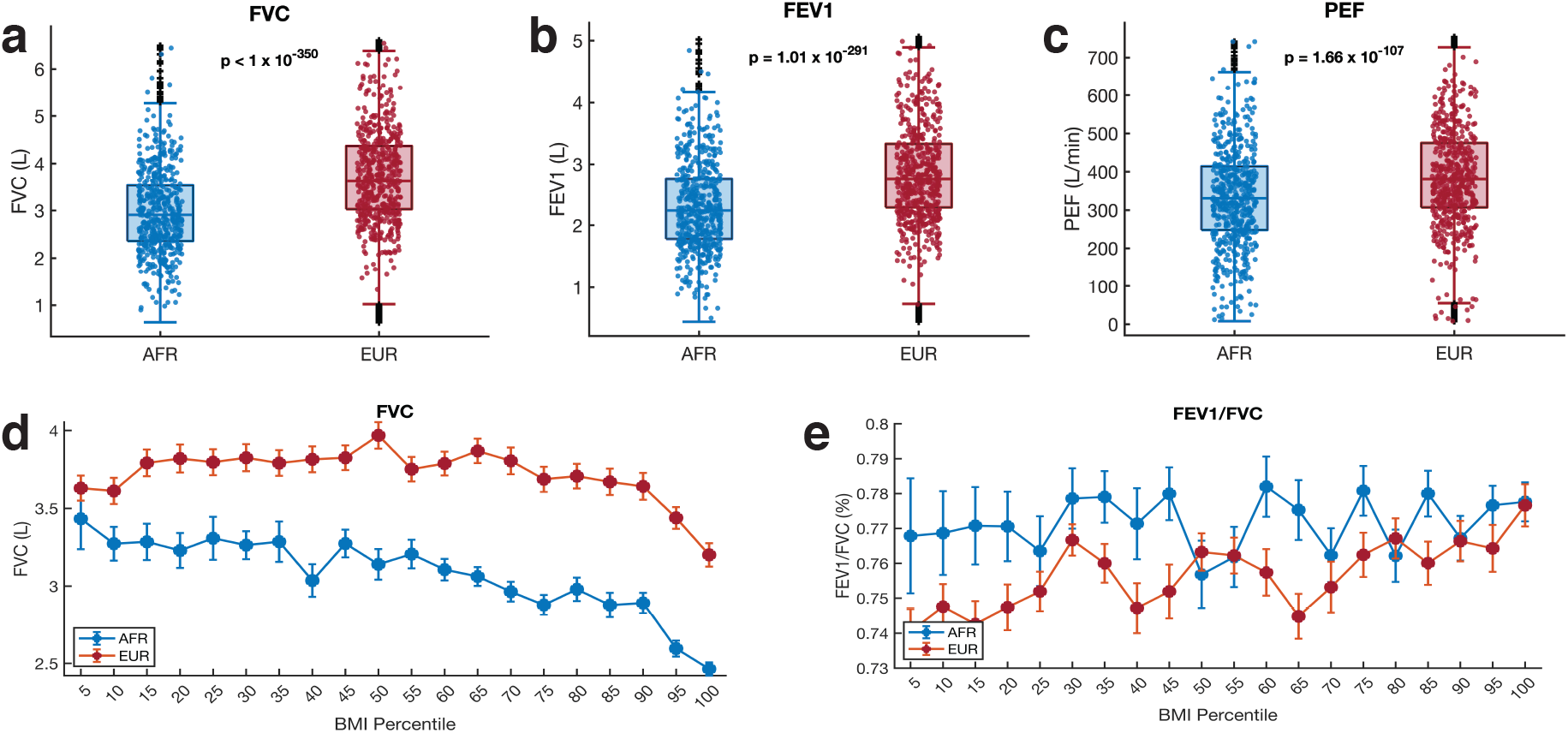
Comparison of the pulmonary function parameter among Africans and Europeans for **(a)** FEV1, **(b)** FVC, and **(c)** PEF. The boxplots indicate the distribution of each test parameter in each group. The p-values shown for each comparison were calculated from Welch’s t-test. On each box, the central mark indicates the median, and the left and right edges of the box indicate the 25th and 75th percentiles, respectively. The whiskers extend to the most extreme data points not considered outliers, and the outliers are plotted individually using the ‘+’ symbol. To make the visualisation clearer, the filled circle mark showing the distribution only include 1000 randomly sampled point from the entire samples size of each group. Error bars showing the variation in **(d)** FVC and **(e)** FEV1/FVC across BMI percentiles among Africans and Europeans. The middle point indicated the mean FVC or FEV1/FVC, and the error bars indicate the standard error of the mean at the particular BMI percentile.

Previous studies show that the FVC, FEV1, and PEF vary with age, body mass index (BMI), and height of the individuals^12–15^. Here, also, we found that FVC, FEV1, and PEF tend to reduce as the age and BMI of the individuals’ increases, and all three parameters increase alongside the height of the individuals (Figure 1d and Supplementary Figure 1a – 1i). Furthermore, we observed that the FVC/FEV1 levels are conversely higher in Africans than Europeans across the BMI percentiles (Figure 1e).

### Genetic variant associated with FVC, FEV1 and PEF among Europeans and Africans

Since the FVC, FEV1 and PEF values were significantly higher in Europeans than in Africans. We presumed that a genome-wide association analysis would identify the genetic variants associated with each of these pulmonary function parameters in each group. Therefore, we collected the GWAS summary statistics for each pulmonary function parameter within each ethnic group (see methods sections). The total number of significant variants (including those in linkage disequilibrium) identified for each pulmonary function parameter is shown in Supplementary Figures 2a, 2b and 2c.

We identified 310 genetic variants significantly associated (p-values < 5 × 10^−8^ and causal probability > 0.1; see methods section) with FVC in Europeans and 2 significant associations in Africans (Figure 2a and Figure 2b). For FEV1, we found 308 significant SNP associations in Europeans and 1 in Africans (Figure 2c and Figure 2d). Furthermore, for PEF, we found 374 significant associations in Europeans and none (0) in Africans (Figure 2e and 2f). Surprisingly, we found that the significant SNPs associated with FVC, FEV1 and PEF for each group were unique (Figure 2g, Figure 2h, Figure 2i and Supplementary File 1).

**Figure 2:**
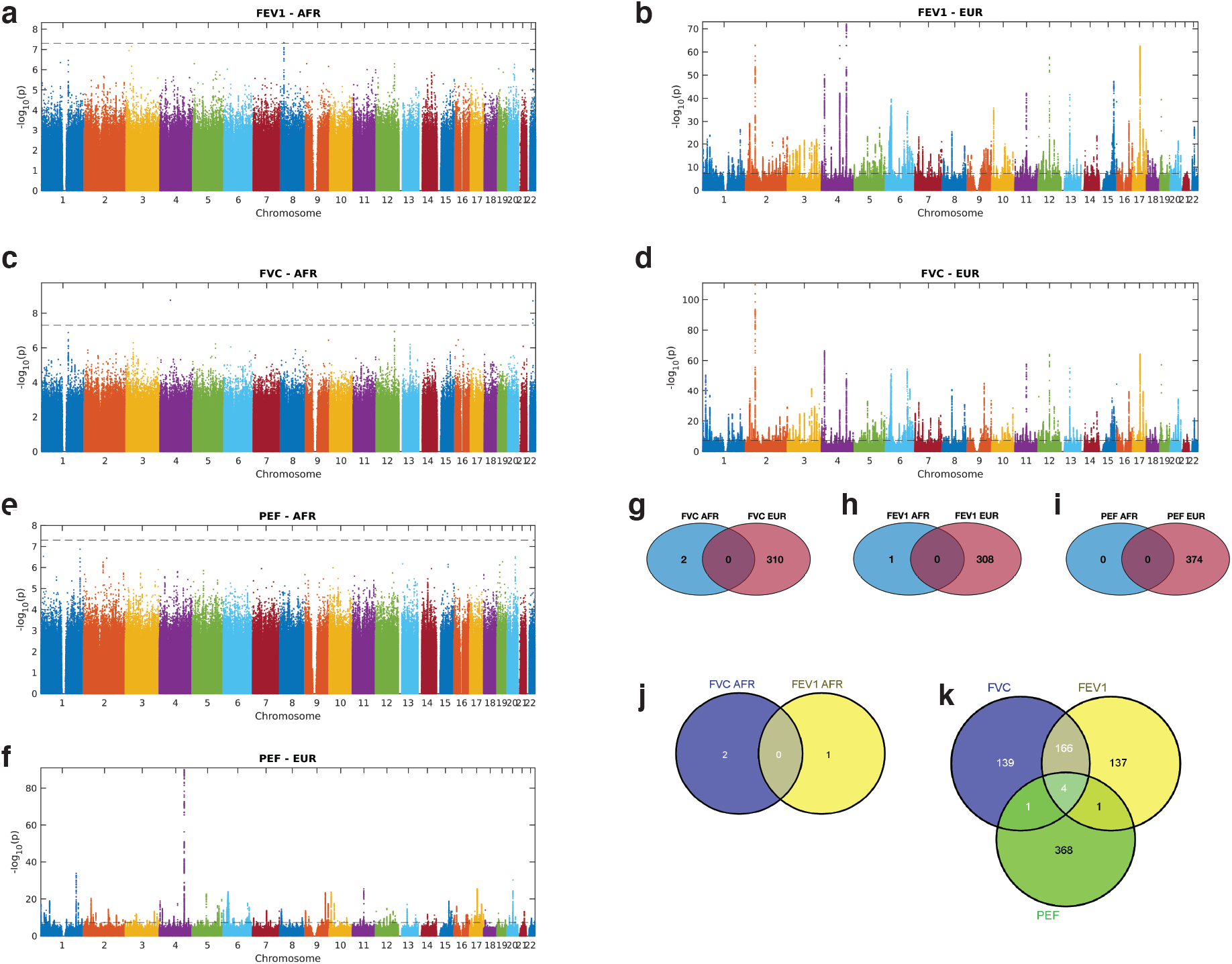
Manhattan plots of the SNPs associated with **(a)** FEV1 in African and **(b)** FEV1 in Europeans, **(c)** FVC in African and **(d)** FVC in Europeans, and **(e)** PEF in African and **(f)** PEF in Europeans for each chromosome. The Venn diagrams show the overlap among the significant causal SNPs associated with **(g)**, FVC **(h)**, PEF, and **(i)** FEV1 in Africans and Europeans. The distribution of genetic variants associated with three pulmonary function parameters among the **(j)** Africans and **(k)** Europeans. Refer to Supplementary File 1 for details concerning individual SNPs and their frequencies among Africans and Europeans.

We compared the 817 unique SNPs in Europeans with the 3 SNPs in Africans significantly associated with the three pulmonary function parameters and found no common variants between the two sets. Conversely, we found that 236 SNPs were associated with FVC and FEV1 in the Europeans (Figure 2k). However, there was no overlap in the associated SNPs among Africans (Figure 2j).

Since the SNPs significantly associated with pulmonary function were unique for Europeans and Africans, we next relaxed the GWAS significance threshold to a suggestive cutoff p-value^16^ of 1 × 10^−6^. Then, we compared the significant SNPs in Europeans and Africans for FVC, FEV1 and PEF. Using a less stringent significance threshold, we still found no common SNPs among Africans and Europeans for all three pulmonary function parameters (Supplementary Figure 2d, 2e and 2f).

We compared the minor allele frequency of SNPs in the UK Biobank between Europeans and Africans for the combined 820 SNPs (817 in Europeans plus 3 in Africans) associated with pulmonary function. We found that 788 out of 820 SNPs differed significantly in their frequency among the African and Europeans (Supplementary File 2). The top-three variants that exhibited the most significant higher frequencies in Europeans compared to Africans were rs2042395 (frequency in EUR = 0.77, in AFR = 0.19, Fisher test p-values = 4.94 × 10^−323^), rs3748400 (EUR = 0.78, AFR = 0.19, p = 6.92 × 10^−323^), rs8045843 (EUR = 0.78, AFR = 0.17, p = 8.89 × 10^−323^), see Supplementary File 2 and Supplementary Figure 2g. Interestingly, the variants rs2042395 and rs8045843 have been previously found to be associated with Well-being spectrum^17^ and Sensitivity to environmental stress and adversity^18^ respectively, in individuals of European ancestry. Conversely, the top variants with higher frequency in Africans compared to Europeans were rs143384 (EUR = 0.40 and AFR = 0.92, p = 2.0 × 10^−323^), rs3133084 (EUR = 0.23 and AFR = 0.65, p = 8.4 × 10^−323^), and rs7853063 (EUR = 0.20 and AFR = 0.60, p = 6.4 × 10^−323^), see Supplementary Figure 2g. Among these, the variant rs143384 has been reported associated with FVC, lung function, and PEF^19^, and among anthropometric traits in Europeans^20^.

Altogether, these analyses revealed that different SNPs may be associated with FVC, FEV1 and PEF among Europeans and Africans and that the frequency of these SNPs significantly varies between these populations.

### Pathway and GWAS Catalogue enrichments of the SNPs

For each study population, we assessed the enrichment of GWAS catalog^21^ annotation terms of the genes in which the SNPs associated (suggestive cutoff p-value of 1 × 10^−6^) with lung function occur (see Supplementary File 2).

The GWAS catalogue term analyses revealed that in Europeans, the genes were significantly enriched for GWAS terms associated with “Height” (combined score [CS] = 1,399, hypergeometric test; p = 1.06 × 10^−93^), “Lung function (FEV1)” (CS = 8291, 5.4 × 10^−25^), “Pulmonary function interaction” (CS = 577.5, p = 2.33 × 10^−19^) among others (Figure 3a, Supplementary File 3). In Africans, we found that the genes were significantly enriched for GWAS terms associated with “Subcutaneous adipose tissue” (CS = 833.6, p = 1.2 × 10^−07^), “Birth weight” (CS = 189.7, p = 3.7 × 10^−04^), “Cognitive decline rate in late mild cognitive impairment” (CS = 18.6, p = 7.3 × 10^−04^), among others (Figure 3b, Supplementary File 3). Overall, these results show that the SNPs identified among Europeans are located in genes known to play roles in many phenotypes, but most notably, those related to pulmonary function or GWAS phenotypes related to pulmonary function. Conversely, the SNPs that we identified associated with pulmonary function among Africans fall within genes that are not enriched for pulmonary function related terms.

**Figure 3:**
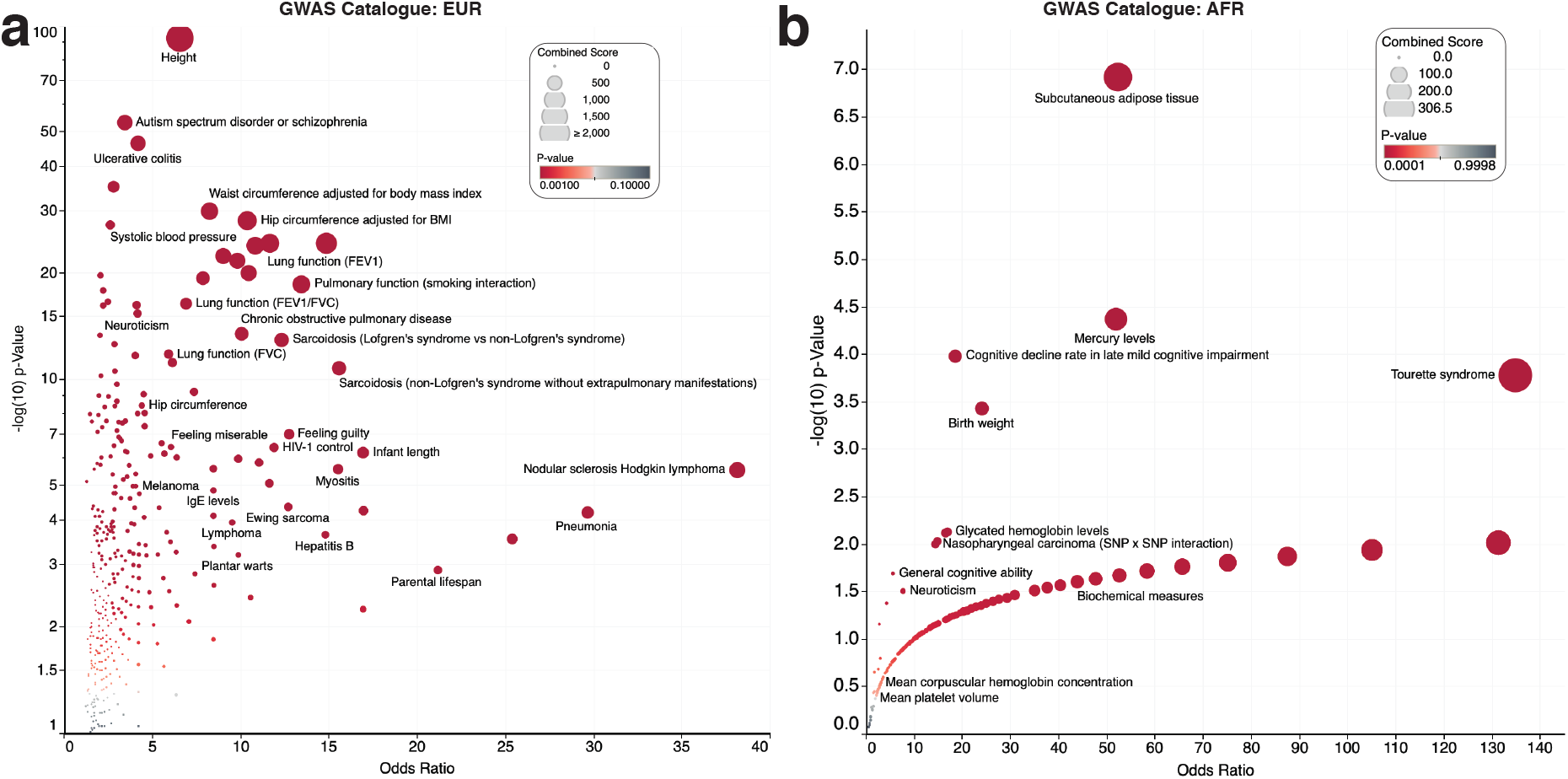
Volcano plots of the GWAS catalogue enrichment analysis for genes in which significant SNPs are located for **(a)** Europeans and **(b)** Africans. All four plots show the adjusted p-value on the y-axis and the odds ratio of the enrichment score on the x-axis. Each circle represents a GWAS catalogue term or Elsevier pathway. The circles are coloured based on the levels of statistical significance, with the redder colours showing a greater degree of significance. Each circle is sized based on the combined enrichment score of the term represented by the circle.

### Variant spanning loci associated with pulmonary function among Europeans and Africans

Many of the associated SNPs may simply reflect the linkage disequilibrium structure of the populations^22,23^ (see Supplementary File 4). For example; we found 10 variants associated with FEV1 and FVC in Europeans within loci 12q14.3, upon fine mapping^24^, found that the most likely causal SNP within the loci was rs1351394 (Probabilistic Identification of Causal SNPs [PICS] value = 0. 7243), a 3-prime UTR variant located in the gene *HMGA2* (Figure 4a). The variant rs1351394 has previously been associated with variations that affect FEV1 capacity, including height ^25,26^ and birth length^27^. Furthermore, *HMGA2* is involved in lung development^28^.

**Figure 4:**
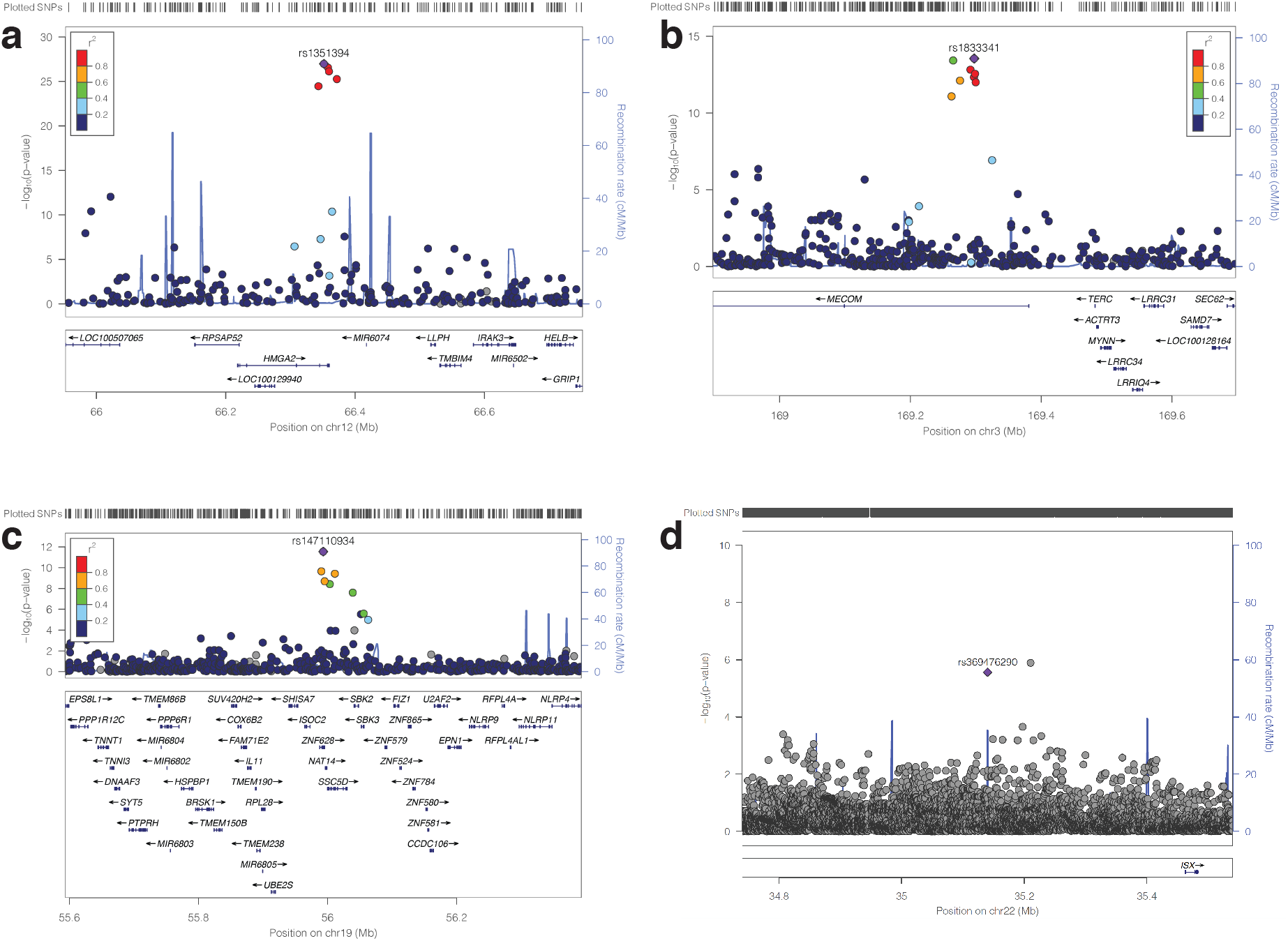
Regional association plots for genome-wide significant pulmonary function loci for the lead SNPs **(a)** rs1351394 at loci 12q14.3, **(b)** rs16909898 at 9q22.32, **(c)** rs147110934 at loci 19q13.42, and **(d)** rs369476290 on chromosome 22. The genes within the chromosomal loci are shown in the lower panel. The blue line indicates the recombination rate. The filled circles show the position of the SNPs along the region on the x-axis and the negative logarithm of the association p-value on the y-axis. The lead SNP is shown in purple, and the SNPs within the locus are coloured based on the linkage disequilibrium correlation value (r2) with the lead SNP based on the European HapMap haplotype from the 1000 genome project.

At the loci 19q13.42, we found that the most likely causal SNP is rs147110934 (PICS = 0.83) associated with both FEV1 and FVC in Europeans (Figure 4b, also see Supplementary File 4). rs147110934 is a predicted missense variant that falls within the *ZNF628* gene. In addition, whilst rs147110934 has not been previously associated with pulmonary function, we found it is associated with height^29^ and body weight^30,31^, both of which are associated with FVC and FEV1.

Furthermore, we found several SNPs in the loci 9q22.32 associated with pulmonary function (Figure 4c). Here, the lead and predicted causal (PICS = 1) variant is rs16909898, located in the *PTCH1* gene previously identified to modify pulmonary function parameters^32,33^ and height^26^.

In addition, for individuals of African ancestry, at the loci 5q32, the lead SNPs among the four associated with pulmonary function was rs369476290 (PICS = 0.67), an intergenic variant located in the gene, *ISX*. rs369476290 has not been previously linked to pulmonary function or disease (Figure 4d).

### Comparison to variants previously associated with pulmonary function

Next, we aimed to identify the previously described and novel SNPs among the significant SNPs that were also predicted causal within a particular LD block (see methods section). Here, we grouped the SNPs into four ordinal categories based on confidence: (1) SNPs reported to be associated with pulmonary function, (2) SNPs related to phenotypes correlated to pulmonary function (e.g., height, see Supplementary Figure 1), (3) SNPs that fall within genes reported to be associated with pulmonary function and/or disease, (4) SNPs that are eQTLs in the lung, and (5) the novels SNPs.

Interestingly, we found that among our list, 85 variants in Europeans and none (0) in Africans have been previously associated with pulmonary function (See Table 1 and Supplementary File 4). These include SNPs in the genes *PLEKHM1, HMGA2, KDM2A*, and *SYTL2* (Table 2). Likewise, we found that 96 variants in Europeans and none (0) of the variants in African ancestry individuals have previously been associated with a phenotype correlated to pulmonary function (see Supplementary File 4). Furthermore, we found that 196 variants in Europeans and 0 variants in Africans are located within genes associated with various pulmonary function phenotypes and disease, and three SNPs in Europeans and none in Africans were significant eQTLs in the lungs. These three SNPs affect the expression of *CAMLG, PHF15* and *MLLT6*. Finally, we found 327 novel SNPs in Europeans and 3 in Africans associated with pulmonary function; see Supplementary File 4 for the complete list of significant variants and the studies reporting the known variants.

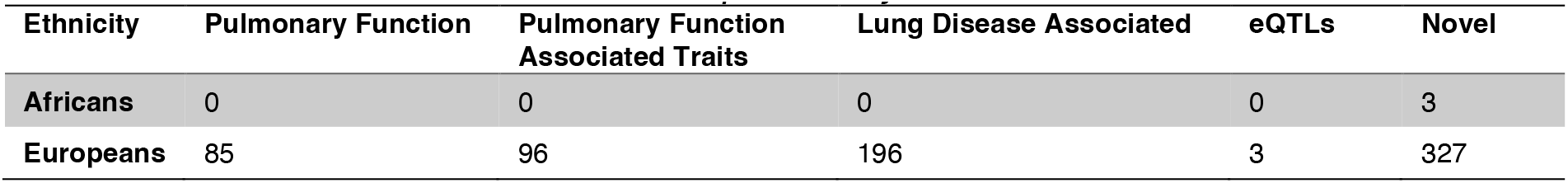
Known and novel variants associated with pulmonary function

Top significant variants associated with pulmonary function

| Variant | Nearest Genes | Ethnicity | Measure | Chrom:Pos | GWAS p | Evidence |
| --- | --- | --- | --- | --- | --- | --- |
| rs536516159 | *LZTS1* | AFR | FEV1 | 8: 20598779 | 4.8 x 10<sup>-08</sup> | Novel |
| rs8756 | *HMGA2, AC090673.2* | EUR | FEV1 | 12: 66359752 | 8.4 x 10<sup>-58</sup> | Pulmonary Function |
| rs55663797 | *PLEKHM1* | EUR | FEV1 | 17: 43544379 | 7.3 x 10<sup>-52</sup> | Lung Disease Assoc. |
| rs1828591 | *HHIP* | EUR | FEV1 | 4:145480780 | 8.3 x 10<sup>-52</sup> | Pulmonary Function |
| rs571481915 | *LPHN3* | AFR | FVC | 4: 61183218 | 1.8 x 10<sup>-09</sup> | Novel |
| rs369476290 | *ISX* | AFR | FVC | 22: 35140134 | 2.0 x 10<sup>-09</sup> | Novel |
| rs8756 | *HMGA2* | EUR | FVC | 12: 66359752 | 1.2 x 10<sup>-63</sup> | Pulmonary Function |
| rs2696624 | *KANSL1* | EUR | FVC | 17: 44326845 | 1.8 x 10<sup>-62</sup> | Novel |
| rs7952436 | *KDM2A, ADRBK1* | EUR | FVC | 11: 67024534 | 9.3 x 10<sup>-58</sup> | Pulmonary Assoc. |
| rs6829956 | *HHIP* | EUR | PEF | 4:145440288 | 1.1 x 10<sup>-90</sup> | Pulmonary Function |
| rs1342062 | *SLC26A9* | EUR | PEF | 1:205912786 | 1.6 x 10<sup>-34</sup> | Lung Disease Assoc. |
| rs143384 | *GDF5* | EUR | PEF | 20: 34025756 | 5.8 x 10<sup>-31</sup> | Pulmonary Function |

We focused on the genes in which the novel SNPs associated with pulmonary function among Europeans located to performed enrichment analyses based on the DisGeNET database^34^, Phenotype and Genotype Integrator database (PheGenI) ^35^. Here, our DisGeNET analysis revealed that the novel genes are enriched for term related to pulmonary function including “Forced expiratory volume function” (p = 9.7 × 10^−13^) and body measures that modify pulmonary function including “Body Height” (p = 1.33 × 10^−15^), see Supplementary Figure 3. Similarly, our PhenGenI enrichment analysis revealed that the genes are enriched for term pulmonary function terms including Forced Expiratory Volume (p = 2 × 10^−4^) and phenotypes associated with pulmonary function including Body Height (p = 4.2 × 10^−07^), see Supplementary Figure 3. Overall, these findings show that despites the SNPs being novel among Europeans, the genes within which the SNPs are located are known to be associated with Pulmonary function.

### Bias in GWAS studies explains why few SNPs were previously associated with pulmonary function in Africans

Since none of the SNPs that we identified associated with pulmonary function among Africans have been reported in the literature, we queried the GWAS catalog^21^ for previous studies of pulmonary function or phenotypes related to pulmonary function (such as asthma) across various ancestry backgrounds. We found those studies to be significantly biased toward individuals of European ancestry (Figure 5a). Also, despite the number of studies conducted on individuals of African ancestry increasing over the last five years, the gap is widening between the number of studies reported on Europeans compared to Africans during the same time interval (Figure 5a). Overall, among the 235 GWAS studies reported on pulmonary function or phenotypes related to pulmonary function, only eight were conducted on Africans/African Americans. In comparison, we found that 120 studies have been conducted exclusively on individuals of European ancestry (Figure 5b). Furthermore, in the same studies, the cumulative samples size of the Europeans in 2021 (10,633,660 individuals) is approximately 235 times greater than that of the Africans (45,189 individuals; see Figure 5c).

**Figure 5:**
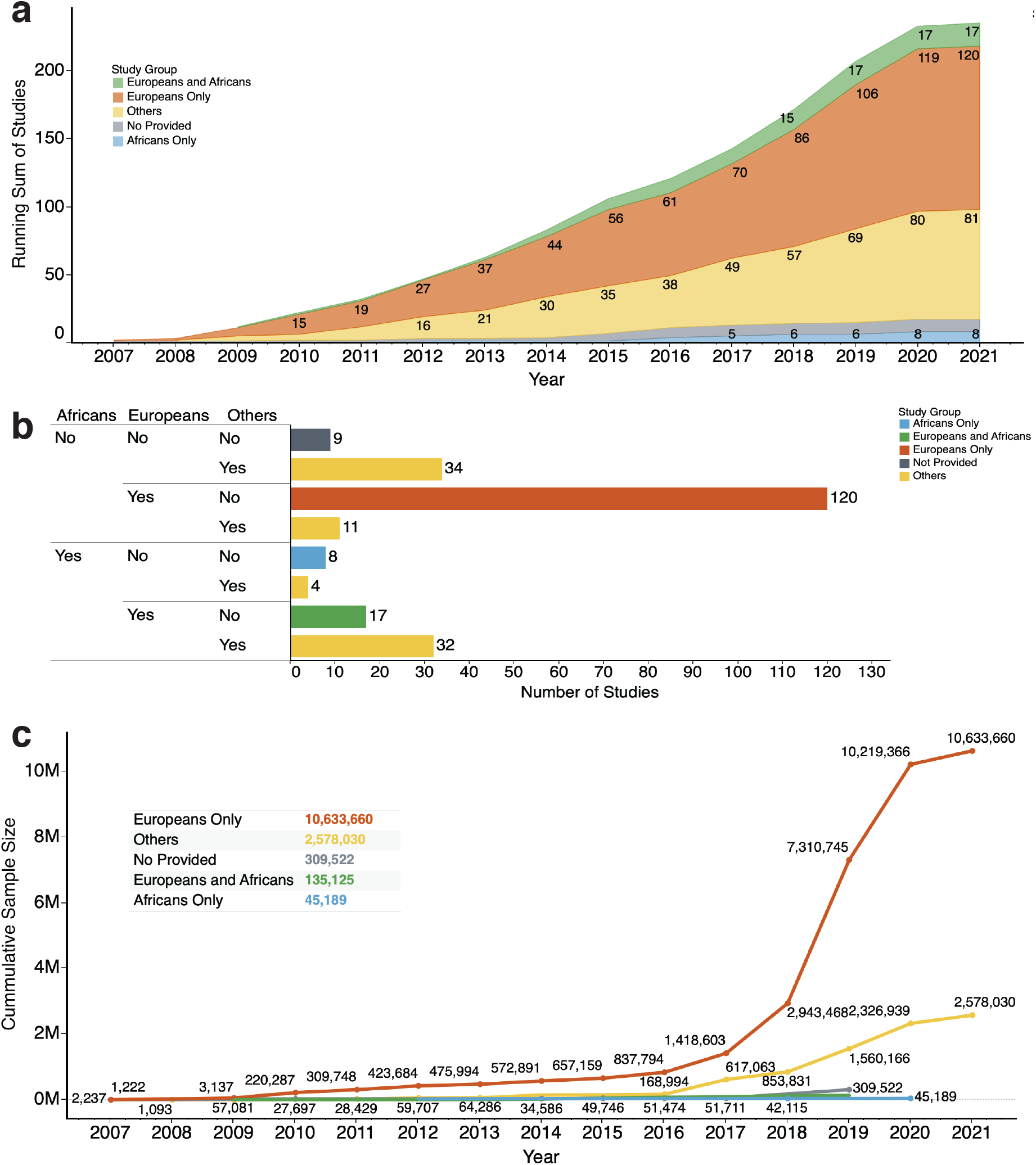
Studies in the GWAS catalogue of pulmonary function and lung phenotypes. **(a)** The plot of the running sum of GWAS studies reported from 2007 to 2021. The colours show details about the race/ancestry groups: Africans only, Europeans only, Europeans and Africans, Others, and those for which the race/ancestry group is “Not provided”. **(b)** The total number of GWAS studies reported for each combination of race/ancestry group. The colours show details about race/ancestry group. **(c)** The trend of the cumulative sum of participants (on the y-axis) of studies from 2007 to 2021. The colours show details about the race/ancestry groups. The marks are labelled by the cumulative sum of participants. The figure insert shows the total number of participants race/ancestry group.

## Discussion

We analysed variations in pulmonary function and the associated genetic variants among individuals of African and European ancestry in the UK Biobank. Here, we report differences in FEV1, FVC, and PEF parameters among Africans and Europeans. Previous studies have examined the pulmonary function parameters between Africans and Europeans, with most reporting the differences we observed ^3,36–38^. However, there has been no explanation for the genetic basis of these observed differences.

Here, we showed that the SNPs associated with pulmonary function were different between Europeans and Africans. Others have reported that the genetic variants associated with various phenotypes may differ among individuals of various ancestry origins^39–42^. This was confirmed by our findings that different variants might be associated with pulmonary function among Africans and Europeans. Despite this observed difference between the two ancestral groups, we are also cognisant that the number of individuals of African ancestry represented in the UK Biobank is much lower than that of Europeans. The smaller sample size of Africans may have resulted in us missing some of the associated SNPs common among the groups^43,44^. It would be interesting to evaluate our findings based on a larger sample of individuals of African ancestry.

Given that the frequency of SNPs, primarily those we found associated with pulmonary function, varies between Africans and Europeans, it is apparent why different variants are associated with these traits^43^. For example, we found that rs12925700 is approximately 21 times more frequent, and rs11205303 is 14 times more frequent in Europeans than Africans, and both SNPs are reported elsewhere^45,46^ and here as being associated with pulmonary function in Europeans. Furthermore, the frequency of genetic variants among individuals of a particular ancestry affects the penetrance of disease and phenotype associated with the alternate alleles ^43,47–50^. Non-alcoholic fatty liver disease^51^, serum uric acid levels^52^, white blood cell count ^53^, fatty acid desaturases^54^, and other phenotypes^55–57^ are associated with different alleles among Africans and Europeans. These alleles are sometimes located on the same gene, but their frequencies vary between ancestral groups.

Our enrichment analyses demonstrated a link between the significant SNPs and GWAS catalogue terms associated with pulmonary function in Europeans, with several of the results showing plausible biological mechanisms. Whereas it was apparent the significantly enriched terms in Europeans were mainly associated with pulmonary function and related phenotypes (Figure 3), we found that the top-ranking terms among SNPs of Africans are not related to pulmonary function. Overall, these results suggest that we need a larger sample size of Africans to identify the variants that modify pulmonary function.

We also showed that genetic association studies of pulmonary function and pulmonary physiology and pathology are significantly biased toward individuals of European ancestry. Even in cases where individuals of African ancestry are included in the studies or studied separately, the number of participants is fewer than those of European ancestry. Furthermore, the trend shows that this gap vis-à-vis how Africans and Europeans are studied has widened over the last few years (see Figure 5).

In summary, we have revealed the extent of variations between Africans and Europeans in the pulmonary function parameters; FEV1, FVC and PEF. In addition, we have identified the different genetic variants associated with pulmonary function among individuals of African and European ancestry. Our integrative analysis of the causal genetic variants, together with the GWAS phenotypes and diseases associated with the genes in which the variants fall, indicate that the significant SNPs are associated with pulmonary function and related phenotypes in Europeans. Therefore, it is apparent that more genetic association studies focused on individuals of African ancestry are required to identify and validate additional causal variants of these traits and other diseases.

## Methods

We analysed a UK Biobank^11^ dataset of 383,471 European (designated as White, British, Irish, and “any other White background”) and 5,978 African ancestries. The demographics of the UK Biobank participants are extensively described elsewhere^11,58^. The data elements that we analysed include genotyping array data of imputed SNPs, anthropometric measurements, and pulmonary function parameters, FVC, FEV1 and PEF.

### Comparison of pulmonary function parameters in Europeans and Africans

We compare the mean values of the pulmonary function parameter FVC, FEV1 and PEF between 383,471 Europeans and 5,978 Africans using the Welch t-test. Furthermore, to evaluate how FVC, FEV1, PEF, and FEV1/FVC values vary with the participant’s body mass index, height, and age, we calculated the 10^th^ percentile bins of each anthropometric measurement and visualised the trend using error bars plotted for each percentile.

#### Genome-wide identification of genetic variants and associations

The methods applied for genotyping of participants in the UK Biobank are reported elsewhere ^11,58^. We obtained the GWAS summary statistics computed by the UK Biobank project of each pulmonary function parameter. The methods used to perform the GWA analyses are described elsewhere ^59^. Briefly, the GWAS was performed for the pulmonary function phenotypes and ancestry groups using the Scalable and Accurate Implementation of GEneralized mixed model approach^60^, using a linear or mixed logistic model including a kinship matrix as a random effect and covariates as fixed effects. The covariates included the participant’s age, sex, age multiplied by sex, the square of the age, the square of the age multiplied by the sex, and the first 10 principal components calculated from the genotype datasets. The Manhattan plots were produced in MATLAB using the software described here^61^. Furthermore, the fine mapping of SNPs to identify the most credible causal SNPs within each linkage disequilibrium block conditioning on the lead SNP signal in each locus ±50 kb was done using the PICS2 software^62^.

### Pathways and Enrichment Analyses

We used NBCI’s dbSNP^63,64^ to ascribe the significant variants associated (suggestive cutoff p-value^16^ of 1 × 10^−6^) with pulmonary function identified using GWAS to specific genes. Then yielded a list of genes associated with pulmonary function in Europeans or Africans. Finally, using these two gene lists (for Europeans and Africans), we separately performed gene set enrichment analysis^65^ using Enrichr^66^ to identify the Elsevier pathways^66^, DisGeNET database^34^, Phenotype and Genotype Integrator database, and GWAS catalog^21^ ontology terms that are significantly enriched for (see Supplementary File 3).

### GWAS literature, disease phenotypes, and eQTLs

We retrieved data of the previous GWAS of pulmonary function and pulmonary function related phenotypes from the GWAS catalog^21^. This information was subset into two categories: “pulmonary reported”; for those studies that reported pulmonary function phenotype and “pulmonary associated” for those that reported associations related to pulmonary function related phenotypes (see Supplementary File 4). In addition, we considered variants as reported when the linkage disequilibrium R-square values of the significant variants that we report here are greater than 0.5 in relation to the previously reported variants elsewhere. Furthermore, we obtained information on diseases associated with the genes in which the variants are located from the Pharos database^67^. Finally, information on SNPs that are eQTLs in the lungs was obtained from the GTEx consortium database^68^.

### Statistics and Reproducibility

We performed the statistical analyses in R programming language, MATLAB 2021a and Bash. We used the Welch test, Wilcoxon rank-sum test and the one-way Analysis of Variance to compare continuous measures among groups. All statistical tests were considered significant if the returned two-sided p-value was < 0.05 for single comparisons. The multiple hypotheses tests were corrected by calculating a two-sided q-value (False Discovery Rate) for each group/comparison using the Benjamini & Hochberg procedure^69^.

## Supporting information

Descriptions of Supplementary Files

## Data Availability

All data produced in the present work are contained in the manuscript

## Data Availability

The datasets that support results presented in this manuscript are available from: the UK Biobank; https://www.ukbiobank.ac.uk and https://pan-ukb-us-east-1.s3.amazonaws.com, dbSNP; https://www.ncbi.nlm.nih.gov/snp, and the GWAS catalogue; https://www.ebi.ac.uk/gwas.

## Code Availability

Code to reproduce most of the results and plots is available from the corresponding authors upon reasonable request.

## Acknowledgements

The funding for this project was provided by H3ABioNet, supported by the National Institutes of Health Common Fund under grant number U24HG006941. The content of this publication is solely the authors’ responsibility and does not necessarily represent the official views of the National Institutes of Health.

## Author Contributions

M.S., S.E., and N.M conceptualized the study. M.S., N.M., S.E., and J.C. designed the methodology, and M.M. M.S, M.M., J.C., and S.E. performed the formal analysis of the data. M.S., N.M., and S.E., drafted manuscript. Editing and reviewing the manuscript was carried out by M.S., N.M., S.E., J.C., and M.M. Data visualisations were produced by M.S. and S.E.

