## Supplementary material for "Genome-wide Association Study of Pulmonary Function in Europeans and Africans from the UK Biobank Identifies Distinct Variants": Descriptions of Supplementary Files

### Description of Additional Supplementary Files

**Supplementary File 1:** Supplementary data of genome-wide associations with pulmonary function. The spreadsheet contains the following results/datasets according to the sheet name. ***FVC AFR:*** SNPs that are significantly associated with FVC in Africans. ***FVC EUR*:** SNPs that are significantly associated with FVC in Europeans. ***FEV1 AFR:*** SNPs that are significantly associated with FEV1 in Africans. ***FEV1 EUR*:** SNPs that are significantly associated with FEV1 in Europeans. ***PEF AFR:*** SNPs that are significantly associated with PEF in Africans. ***PEF EUR*:** SNPs significantly associated with PEF in Europeans.

**Supplementary File 2:** Frequency of genome-wide associated SNPs with pulmonary function among Africans and Europeans: The spreadsheet contains the following results/datasets according to the sheet name. ***SNP Freq – Annon;*** Frequency of SNPs in Africans and Europeans and the related p-value of the frequency difference calculated using the Fisher exact test. The sheet also contains information on which the phenotype(s) associated with the particular SNP. ***Location of SNPs;*** Gene within which the significant SNPs are located for Africans and Europeans.

**Supplementary File 3:** GWAS enrichment analyses: The spreadsheet contains the following results/datasets according to the sheet name. ***GWAS Catalog-EUR;*** GWAS catalogue terms that we found significantly enriched in Europeans based on the genes in which the genome-wide significant SNPs associated with pulmonary function are located. ***GWAS Catalog-AFR;*** GWAS catalogue terms that we found significantly enriched in Africans based on the genes in which the genome-wide significant SNPs associated with pulmonary function are located.

**Supplementary File 4:** Classification of causal variants associated with pulmonary function: The spreadsheet contains the following results/datasets according to the sheet name. ***PulmonaryReported;*** SNPs reported in GWAS catalogue to be associated with pulmonary function. ***PulmonaryAssociated;*** SNPs related to phenotypes correlated to pulmonary function (e.g., height, see Supplementary Figure 1). ***LungDiseaseAssociated;*** SNPs that fall within genes reported to be associated with pulmonary function and/or disease. ***eQTL;*** SNPs that are eQTLs in the lung as reported by the GTEx project. ***Novel;*** the novels SNPs, i.e., that do not meet any of the previously listed criteria. ***Lead SNPs;*** the top-lead SNPs that are found associated with pulmonary function in Europeans and Africans.

**Supplementary File 5:** List of GWAS catalogue studies of pulmonary function and related phenotypes from 2007 to 2022 on which Figure 5 is based.
