## Supplementary Figures for "Genome-wide Association Study of Pulmonary Function in Europeans and Africans from the UK Biobank Identifies Distinct Variants"

### Supplemental Information Titles and Legends

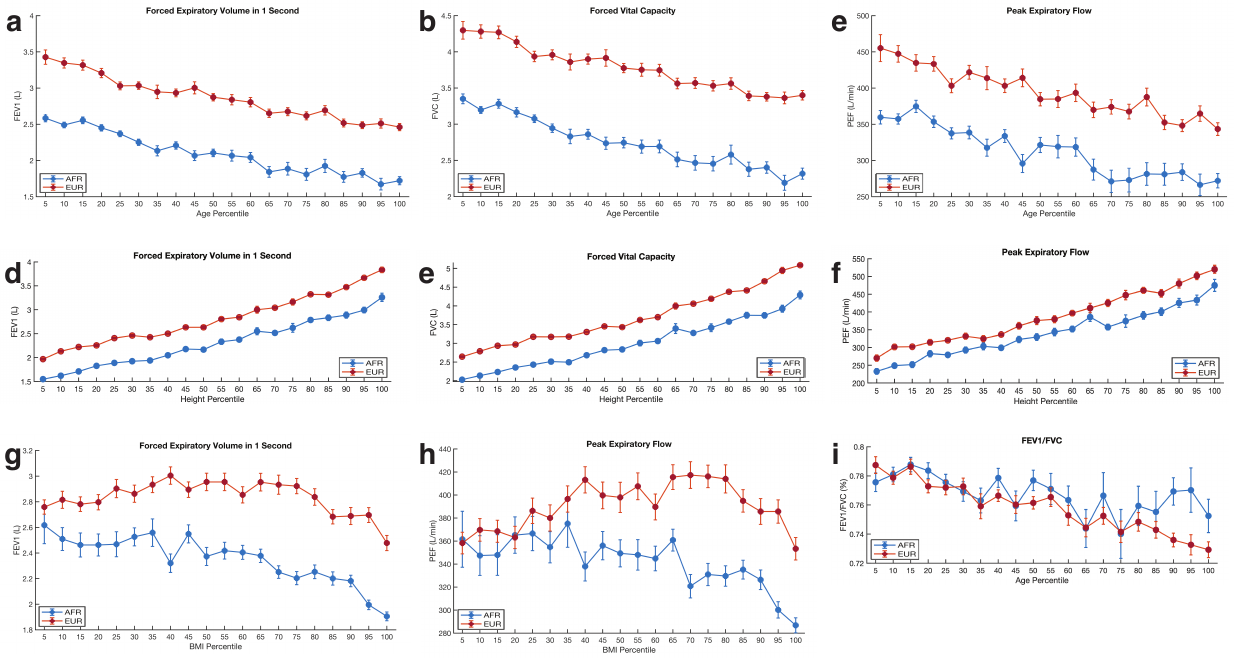

***Supplementary Figure 1:*** *Relationship between Pulmonary function parameters to some anthropometric measurements.* *The error bars show the variation in* ***(a)*** *FEV1,* ***(b)*** *FVC and* ***(c)*** *PEF across age percentiles among Africans and Europeans. Variations in* ***(e)*** *FEV1,* ***(f)*** *FVC and* ***(g)*** *PEF across height percentiles among Africans and Europeans.* ***(h)*** *FEV1 and* ***(i)*** *PEF across BMI percentiles and* ***(j)*** *FEV1/FVC across age percentiles. The middle point indicates the mean parameter value, and the error bars indicate the standard error of the mean at the particular height, age, or BMI percentile.*

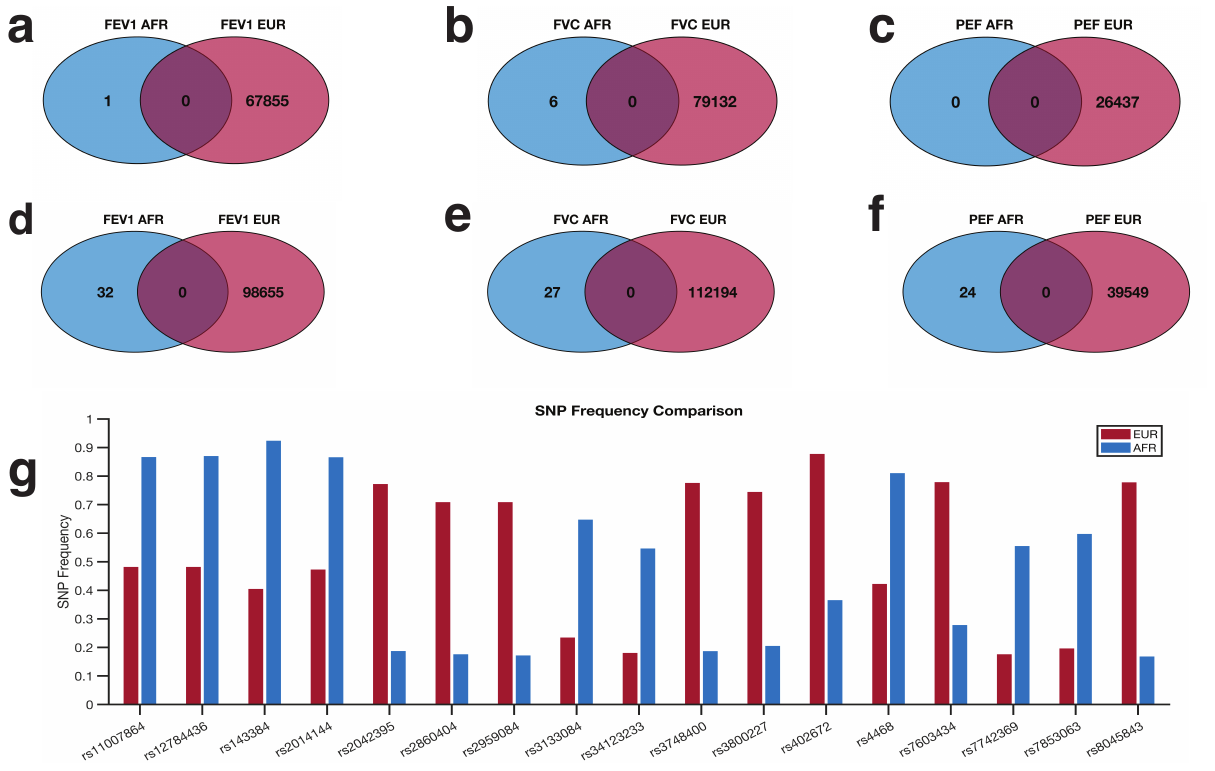

***Supplementary Figure 2:*** *The Venn diagrams showing the distribution of genetic variants the significant (p-value of 5 x 10^-8^, irrespective of the linkage disequilibrium and causal probability) variants associated with* ***(a)****, FEV1* ***(b)****, FVC****,*** *and* ***(e)*** *PEF in Africans and Europeans. The Venn diagrams show the distribution of genetic variants the significant SNPs associated with* ***(d)****, FEV1* ***(e)****, FVC****,*** *and* ***(f)*** *PEF in Africans and Europeans based on the suggestive p-value of 1 x 10^-6^.* ***(g)*** *Bar graph showing the SNPs associated with pulmonary function that exhibit the most significant difference in frequencies among African and Europeans. Refer to Supplementary File 1 for details concerning individual SNPs and their frequencies among Africans and Europeans.*

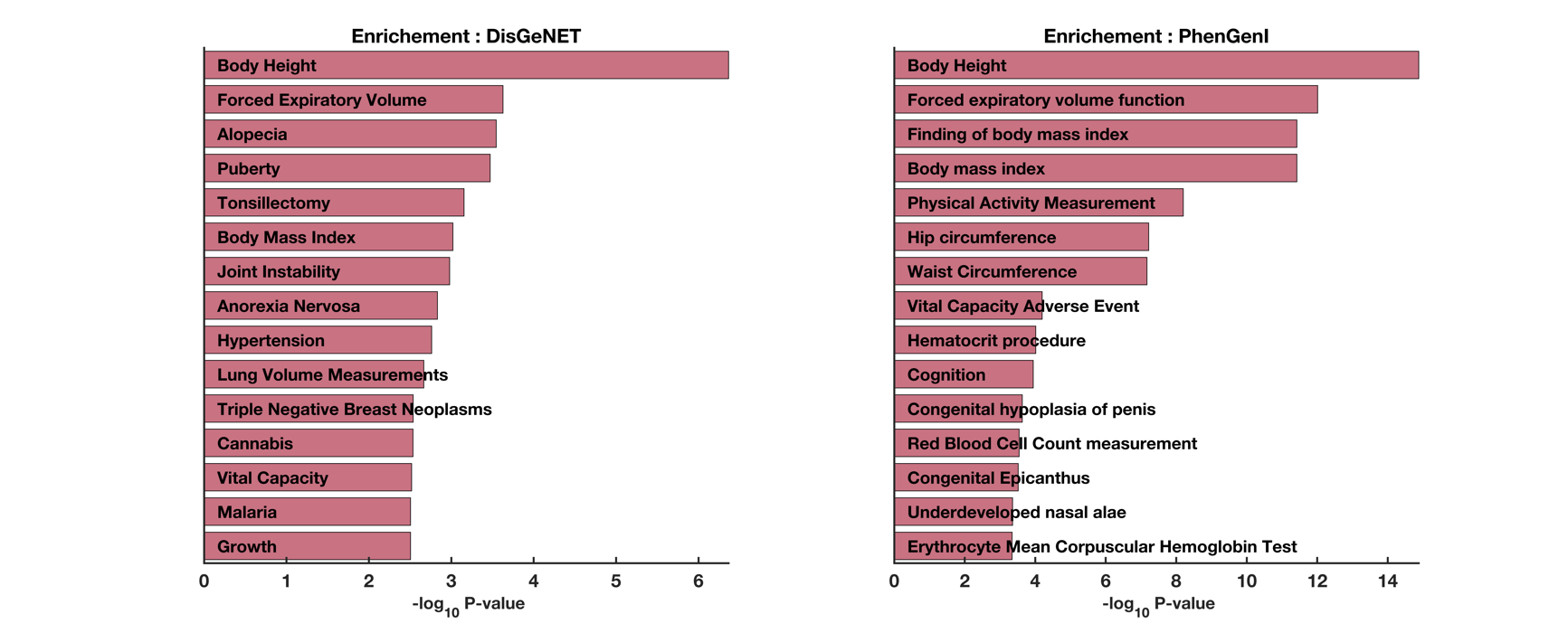

***Supplementary Figure 3:*** *Enrichment analysis results using the genes in which the novel SNPs are located based on the DisGeNET database and PheGenI database.*
